# Evaluation of a novel respiratory virus inactivating buffer for parallel RT-qPCR and quick antigen testing

**DOI:** 10.1101/2024.03.06.24303861

**Authors:** Sigrid Deprez, Sophia Steyaert, Mikhail Claeys, Ludwig Verhasselt, Jo Vandesompele

## Abstract

Since the COVID-19 pandemic, many hospitals implemented a dual testing procedure for SARS-CoV-2 to assess the infection risk of an admitted patient. To allow for a short turn-around time, rapid antigen (Ag) testing via lateral flow tests (LFT) is combined with nucleic acid amplification testing (NAAT), requiring two nasopharyngeal swab collections. In this study, a novel, universal pathogen inactivation buffer (DNA/RNA Defend Pro (DRDP)) was evaluated for SARS-CoV-2 inactivation and simultaneous Ag and DNA/RNA stabilization.

In an emergency department setting of a General Hospital in Ghent (Belgium), patients were tested for SARS-CoV-2, whereby a LFT for Ag detection (Abbott Panbio COVID-19 Ag Rapid Test) was performed in combination with sample collection for NAAT (Abbott Alinity m). Left-over buffers from LFT were diluted in DRDP to evaluate LFT and NAAT results after dilution. Thirty-six patients were included in the data analysis.

Twenty-three diagnostic LFT results were available in the laboratory information system of which all corresponded with results after dilution with DRDP. When correlating NAAT results, seven out of eight positive test results were in agreement, compared to twenty-three out of twenty-five negative results. For thirty-four out of thirty-six samples, LFT and NAAT after dilution with DRDP yielded the same conclusion. Additionally, RNA stability in DRDP was demonstrated when stored for three days at room temperature. At the extreme, a sample stored in the DRDP buffer for 53 days at room temperature was still very positive (Cq 21.95).

We demonstrated that, for the first time, a novel collection buffer could inactivate a pathogen (SARS-CoV-2) while also preserving antigen (for rapid antigen testing) and RNA (for molecular testing). This novel buffer holds promise for a single specimen to be used for both antigen and molecular testing in a safe working environment.

## 1. INTRODUCTION

In 2019, a new coronavirus, severe acute respiratory syndrome coronavirus-2 (SARS-CoV-2), emerged from Wuhan (China) causing a worldwide pandemic ^1^. SARS-CoV-2 causes severe disease to human known as coronavirus disease 2019 (COVID-19). This pandemic illustrated the importance of diagnostic tests to identify SARS-CoV-2 infections and thereby allowing to slow down the spread of the virus ^2^. The need for diagnostic tests led to the fast development of testing methods including rapid antigen (Ag) testing based on lateral flow tests (LFT) and nucleic acid amplification testing (NAAT) using polymerase chain reaction (PCR)-based tests ^3^. From a biosafety perspective, virus inactivating protocols were needed to permit save transportation of diagnostic sampling material for NAAT in dedicated laboratories. Inactivating buffers have recently been developed for that purpose, allowing for safe NAAT, simultaneously inactivating and preserving clinical samples. An example of such a pathogen inactivating, RNA/DNA stabilizing transport medium for in vitro diagnostic testing using PCR methods is the InActiv Blue medium (InActiv Blue, Beernem, Belgium) ^4–9^.

In healthcare institutions in Belgium and abroad, two-step protocols have been introduced during different SARS-CoV-2 waves to allow for screening and confirmation of infected individuals. At the Maria Middelares General Hospital (Ghent, Belgium), an initial screening at the emergency department (ED) is performed using a rapid Ag test, allowing for a very short turn-around time (TAT) of only 15 minutes between sampling and result. The rapid Ag test result is then followed by confirmation by NAAT. Such a two-step protocol allows for immediate action and isolation in case of a positive test, while gaining insight in the viral load based on the NAAT result. However, a disadvantage is the need for sampling of two separate nasopharyngeal swabs because of the absence of a universal medium. A major drawback of most commercially available, pathogen inactivating transportation media, is their limited Ag stabilizing effect while being suited for NAAT testing. Vice versa, Ag testing buffers mostly do not have RNA/DNA stabilizing properties, nor pathogen inactivation properties. Currently, at AZ Maria Middelares, nasopharyngeal swabs are collected in InActiv Blue (InActiv Blue, Beernem, Belgium) inactivation medium allowing safe sample handling and transportation ^4^.

Through the development of more universal media, stabilizing RNA while also keeping Ag intact, combined testing on one sample would become possible. After performing NAAT, also other Ag tests for respiratory pathogens could be conducted. This is a potential benefit compared to classic, commercial inactivation media. InActiv Blue recently developed such a “universal medium”, the DNA/RNA Defend Pro medium (DRDP) for combined Ag and PCR testing ^10^.

A similar medium has been developed by RNAssist, “virusPHIX” and “virusPHIX-P9” (Rapid Labs, Essex, England) in 2020 ^11^. The VirusPHIX medium has been developed for virus inactivation and RNA stabilization, offering safe collection and transport of SARS-CoV-2 clinical swab and saliva samples to clinical laboratories for both COVID-19 LFT and PCR testing. The medium has been developed for research use only. Nevertheless, no scientific literature has been published demonstrating the effectiveness of the medium.

The primary objective of this study was to evaluate the performance of the DRDP medium for SARS-CoV-2 rapid Ag testing as well as its suitability for SARS-CoV-2 NAAT testing compared to the currently used protocols for SARS-CoV-2 Ag and NAAT testing at the clinical laboratory of General Hospital AZ Maria Middelares. Additionally, for samples positive for other respiratory viruses (Influenza A and B, and respiratory syncytial virus (RSV)), preliminary data was collected on the performance of DRDP for NAAT testing in comparison with InActiv Blue (current diagnostic medium).

## 2. MATERIALS AND METHODS

### 2.1 Sample collection and analysis

Left-over buffers of LFT for SARS-CoV-2 antigen (Ag) testing, sampled from patients admitted at the ED of the Maria Middelares General Hospital (Ghent, Belgium) over a period from January 2023 until April 2023, were collected. Initially, an inclusion target of 50 patients was set. This study was approved by the Ethics Committee of Maria Middelares General Hospital (EC MMS.2023.045).

As part of the standard clinical care in the hospital, two distinct nasopharyngeal swabs were sampled of admitted patients at the ED to determine the SARS-CoV-2 infection risk, as depicted in **Figure 1**.

**Figure 1.**
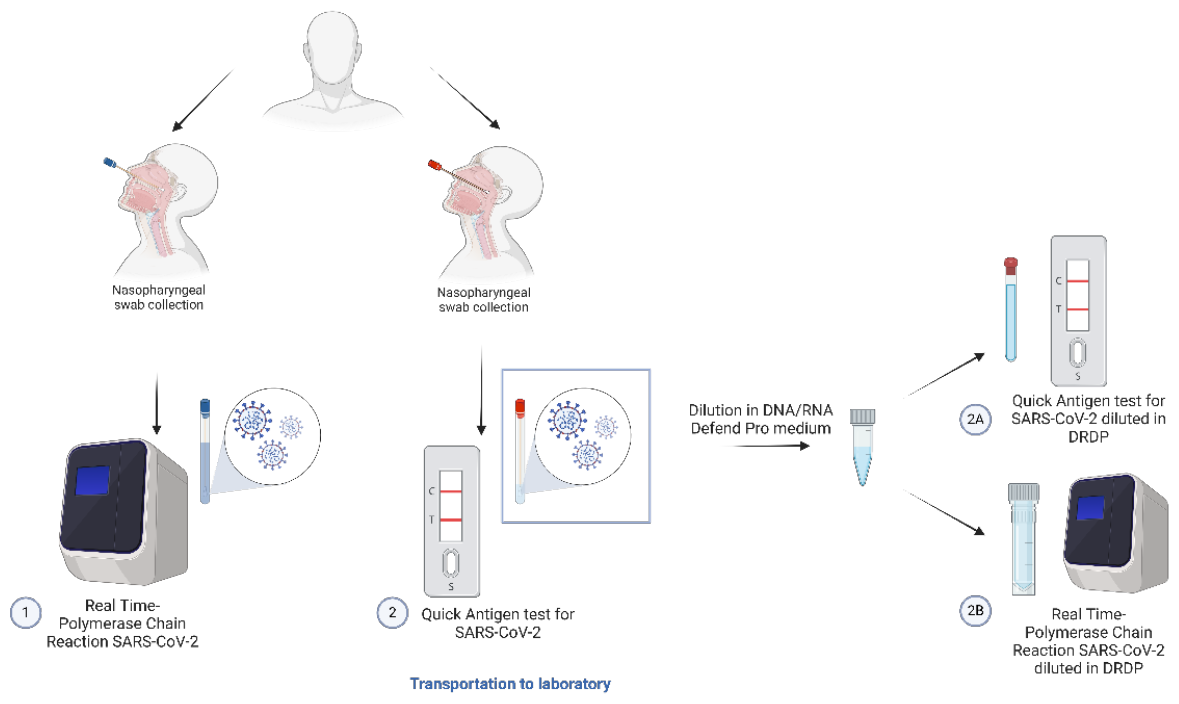
Graphical representation of sample collection of a patient at the emergency department and further processing in the laboratory followed by dilution in DNA/RNA Defend Pro medium (DRDP).

Using a first nasopharyngeal swab (red swab depicted in **Figure 1**), a LFT to detect SARS-CoV-2 Ag was performed at the ED by trained medical personal, using Abbott Panbio COVID-19 Ag Rapid Test Devices (Abbott, Chicago, Illinois, United States) in accordance with the product leaflet ^12^. Additionally, a second swab was collected (blue swab depicted in **Figure 1**) in virus inactivating medium (InActiv Blue, Beernem, Belgium) for save transportation to the laboratory for NAAT for SARS-CoV-2. As outlined above, InActiv Blue (IAB) is a virus inactivation and RNA-stabilizing lysis buffer ^4^. Upon arrival at the microbiology laboratory, NAAT was performed by real time reverse transcriptase polymerase chain reaction (RT-PCR), conducted on an Abbott Alinity m platform (Abbott, Chicago, Illinois, United States). Multiplex Resp-4-Plex assays were used to simultaneously detect SARS-CoV-2 virus, Influenza A and B, and Respiratory Syncytial Virus (RSV) given the flu season at the time of conducting the study ^13^.

Within the framework of this study, left-over sample buffer in test tubes and test cartridges of Panbio COVID-19 Ag Rapid Test Devices (test number 2 at **Figure 1**) were transported in a biohazard safety bag to the clinical laboratory at the Maria Middelares General Hospital using pneumatic transportation tubes. Upon arrival at the laboratory, buffer tubes and test cartridges were pseudonymized with a patient study code (COV-xx). A photographic image was taken of the original test cartridges when available (test number 2 in **Figure 1**) for visual comparison with the test cartridge result after sample buffer dilution in DRDP (test number 2A in **Figure 1**), as outlined below.

Following pseudonymization of the buffer tubes and cartridges, left-over samples were diluted in DRDP. A minimal volume of 50 µL from the left-over sample was diluted to a total volume of 350 µL with DRDP medium in an Eppendorf tube. This was followed by a vortexing and homogenization step. From the Eppendorf tube, 250 µL was subsequently transferred to a new test tube (Abbott rapid antigen test kit). A fixed volume of base (1 M NaOH, 6.8% v/v, equi) was added to the new test tube to bring the pH to > 6.5 (i.e. 16-17 µL). An acidic pH would precipitate the colloidal gold in the Ag test device during analysis. After vortexing the tube, a second LFT (Panbio COVID-19 Ag rapid test) was performed. The analysis was performed as prescribed on the leaflet. No additional nasopharyngeal sampling was required, since the buffer tube was filled with sample diluted in DRDP (see above). Five drops of sample were dispensed vertically into the specimen well on the cartridge device. After 15 minutes (not more than 20 minutes) the results were read from the cartridge. A photographic image was taken when the results were interpreted for visual comparison with the original (diagnostic) test cartridge result.

Secondly, the remaining 100 µL in the Eppendorf tube was transferred to a tube for PCR-analysis, as depicted in **Figure 1**, test 2B. The left-over buffer was further diluted with DRDP to a total volume of 1250 µL (1500 µL for some positive samples because of a smaller left-over volume for reanalysis). Here, no 6.8% (v/v) basic solution was added for performance of NAAT.

Samples in DRDP that tested positive for SARS-CoV-2 with NAAT (PCR tubes), were stored for 3 days at room temperature for reanalysis to assess sample stability in DRDP.

### 2.2 Method comparison

LFT results for SARS-CoV-2 were interpreted as ‘negative’, ‘positive’ or ‘invalid’ in agreement with the product leaflet ^12^. Rapid Ag test results as part of diagnostic care (**Figure 1**, test 2) were compared with test results after dilution in DRDP (**Figure 1**, test 2A). Agreement between paired samples was evaluated.

Using Abbott Alinity m NAAT (Resp-4-Plex), Cq values were obtained for the detection of SARS-CoV-2, Influenza A, Influenza B, and RSV DNA/RNA. Based on the SARS-CoV-2 Cq value, samples were categorized as ‘negative’ (no detection), ‘weak positive’ (Cq > 24.82), ‘positive’ (18.43 < Cq ≤ 24.82), ‘strong positive’ (12.04 < Cq ≤ 18.43) and ‘very strong positive’ (Cq ≤ 12.04). Categorical NAAT results as part of diagnostic care (**Figure 1**, test 1) were compared with NAAT results after dilution in DRDP (**Figure 1**, test 2B). Importantly, paired NAAT results originate from different samplings, as visually indicated in **Figure 1**. Besides the comparison of categorical results between paired NAAT samples, delta Cq values were calculated. Using delta Cq values (Δ Cq), the dilution factor in DRDP can be taken into account. Additionally, it allows us to compare the results across different samples.

For each sample, Cq values obtained as part of diagnostic care (sampled in InActiv Blue, IAB), were subtracted from Cq values obtained after dilution in DRDP to obtain a Δ Cq value (Δ Cq = Cq DRDP – Cq IAB). After calculating Δ Cq values for each sample, a mean and median Δ Cq value for SARS-CoV-2 for all samples was compared to the expected Δ Cq based on sample dilution. For NAAT, samples were

87.5 times diluted in DRDP (**Figure 1**, test 2B) when considering a minimal left-over volume of 50 µL. A theoretical Δ Cq of 6.45 was thus expected (Δ Cq_expected_ = log_2_(87.5)). To allow for a more accurate estimation of expected Δ Cq, left-over sample volumes were measured approximately and taken into account for calculation of expected Δ Cq. Again, note that the original Cq and diluted Cq come from an independent swab.

Additionally, Δ Cq values for SARS-CoV-2 were calculated for positive DRDP samples analyzed on day 0 and day 3 (Δ Cq_stability_ = Cq DRDP day 3 – Cq DRDP day 0), and mean and median Δ Cq values were determined. Here a Δ Cq_expected_ of zero was expected, since no additional dilution was performed and RNA stability assumed.

Thirdly, LFT results in DRDP (**Figure 1**, test 2A) and DRDP SARS-CoV-2 NAAT results (**Figure 1**, test 2B) were compared qualitatively (‘positive’ or ‘negative’) to assess agreement between paired samples in DRDP, originating from the same sampling.

Finally, for samples positive for Influenza A, Influenza B, or RSV, Δ Cq values were also determined and compared to the expected Δ Cq (Δ Cq_expected_ = log_2_(87.5) = 6.45), as outlined above. Also here, to allow for a more accurate estimation of expected Δ Cq, left-over sample volumes were measured approximately and taken into account for calculation of expected Δ Cq. Also here, the original Cq and diluted Cq come from an independent swab.

## 3. RESULTS

### 3.1 Lateral flow test results – SARS-CoV-2

In total, 36 left-over samples were collected. For 23 out of 36 samples, LFT results were available in the laboratory information system (LIS). Seven samples (on a total of 23) were reported positive, sixteen as negative (**Table 1, “Panbio”**). For all seven positive samples, concordant positive results were obtained after dilution in DRDP, as displayed in **Table 1, “DRDP”**. All sixteen negative samples also tested negative after dilution. For one of the remaining 13 samples, for which a negative result was mentioned in the electronic patient file, a weak positive result was obtained after dilution. However, no official report was received by the laboratory (not included in **Table 1** since no LIS result was reported). For the remaining twelve samples, although performed at the ED, no diagnostic LFT result was reported.

**Table 1.** Results LFT for SARS-CoV-2 (Diagnostic Panbio vs. after dilution in DRDP)

|  | positive Panbio | negative Panbio | total |
| --- | --- | --- | --- |
| positive DRDP | 7 | 0 | 7 |
| negative DRDP | 0 | 16 | 16 |
| total | 7 | 16 | 23 |

Additionally, intensities of LFT results were compared, as depicted in **Figure 2**. Although no quantitative result can be deducted from a LFT, Ag-band intensities for SARS-CoV-2 correspond quite well for diagnostic LFT versus LFT after dilution in DRDP. Overall, comparing Ag testing results before (diagnostic test) and after dilution (in DRDP), all results were in agreement. The dilution in DRDP with a median factor of 5.0 (range 1.9-9.3), did not significantly influence LFT results nor the test sensitivity or specificity compared to the diagnostic LFT.

**Figure 2.**
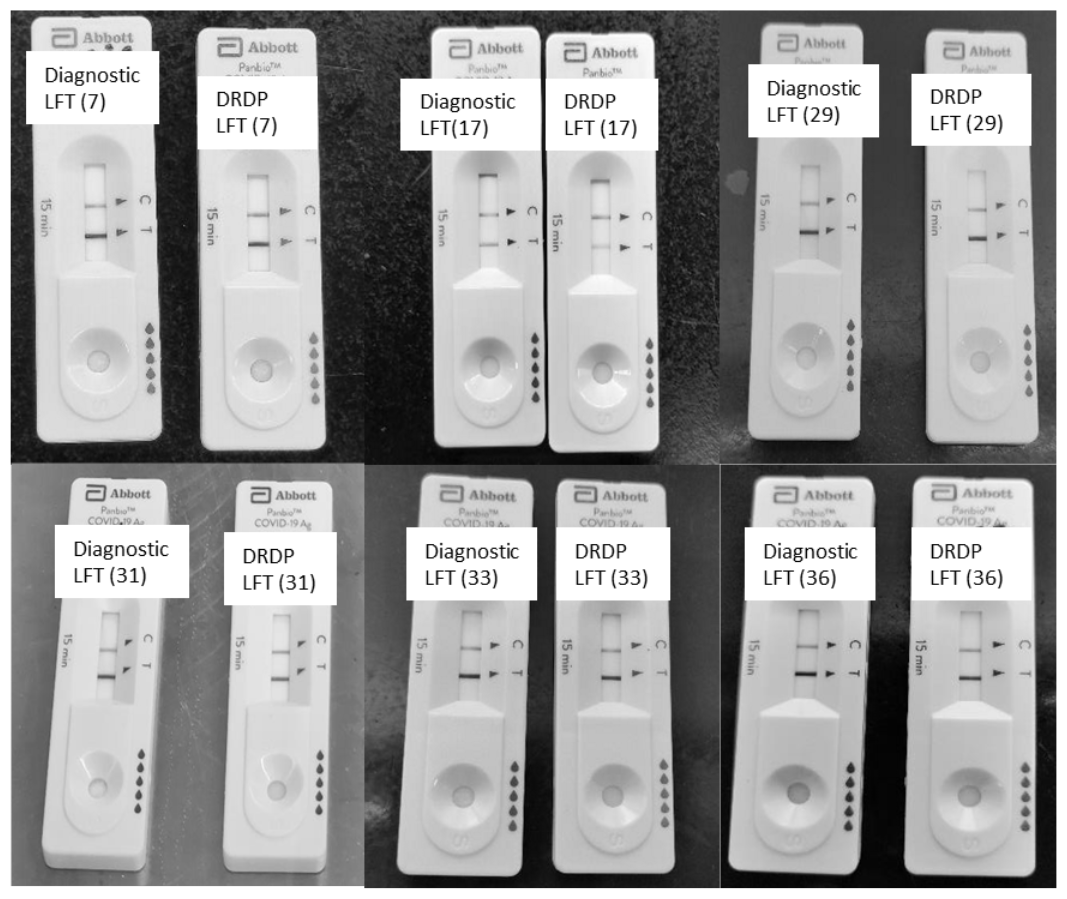
Comparison of SARS-CoV-2 Ag detection intensities for six samples on diagnostic LFT vs. LFT after dilution in DRDP. Numbers between brackets reflex sample numbers as indicated in Supplementary Data File 1.

### 3.2 Nucleic acid amplification test results – SARS-CoV-2

For three out of 36 samples, no NAAT results in IAB buffer were available in the LIS. For 8 samples, Cq values were obtained ranging from 11.28 to 23.31. No SARS-CoV-2 RNA was detected in 25 samples (**Table 2**). Using the left-over LFT samples, after dilution in DRDP, 7 of 8 NAAT positive samples tested positive with Cq values ranging from 14.70 to 33.23. One sample negativized after dilution of the LFT left-over buffer, although a diagnostic Cq value of 17.69 (categorized as strong positive) was obtained. Comparison with diagnostic LFT results (as outlined below) was used to clarify this discrepancy. Importantly, the diagnostic test results compared with NAAT results after dilution in DRDP originate from two different nasopharyngeal swab samplings.

**Table 2.** Results molecular testing SARS-CoV-2 (diagnostic Abbott Alinity m vs. after dilution of LFT sample in DRDP)

|  | positive InActiv Blue | negative InActiv Blue | total |
| --- | --- | --- | --- |
| positive DRDP | 7 | 2 | 9 |
| negative DRDP | 1 | 23 | 24 |
| total | 8 | 25 | 33 |

**Table 3.** Results molecular testing SARS-CoV-2 compared to LFT for SARS-CoV-2 after dilution in DRDP.

|  | positive LFT DRDP | negative LFT DRDP | total |
| --- | --- | --- | --- |
| positive NAAT DRDP | 9 | 2 | 11 |
| negative NAAT DRDP | 0 | 25 | 25 |
| total | 9 | 27 | 36 |

When comparing categorical NAAT results, which are important for test interpretation and management in the hospital, for the 8 positive NAAT samples in IAB, results in DRDP differed for 6 samples with not more than 1 category, for 1 sample with 2 categories (strong positive vs. weak positive), and 1 sample tested negative in DRDP.

Δ Cq values values between NAAT in IAB buffer (diagnostic PCR test) and in DRDP ranged from 1.33 to 20.64, with a mean and median Δ Cq of 5.77 and 3.28 for SARS-CoV-2. These values are in close agreement with Δ Cq_expected_ value of 5.64 (1/50 dilution) despite the fact that different samplings are compared. Δ Cq_expected_ was calculated based on the exact sample dilutions as the left-over sample volumes ranged from 40-200 µL. Detailed calculations can be found in **Supplementary Data File 1**. For two samples, no SARS-CoV-2 RNA was detected with the diagnostic test in IAB, and a positive result was obtained after dilution in DRDP. In both instances, a weak positive test result was obtained (Cq values of 38.26 and 37.47).

Additionally, NAAT (DRDP) results were compared when analysed on day 0 and day 3 after storage at room temperature. Categorical NAAT results differed maximally with 1 category between paired results. Absolute Δ Cq values ranged from 0.38 to 1.29 with a mean and median Δ Cq of 0.24 and 0.43, indicating no stability issues occurred when stored at room temperature. One sample was even stored for 53 days at room temperature for which a Cq value of 21.95 was obtained. Although no test result was available at day 0, the fact that still a ‘positive’ test result could be obtained indicates a good stabilization of viral RNA at room temperature for prolonged time. Similar stability results were obtained for saliva collected in DRDP (1.2 mL saliva in 2 mL DRDP), whereby delta Cq was < 1 when stored for 2 weeks at room temperature (data not shown) ^10^.

### 3.3 Comparison of lateral flow and nucleic acid amplification test results – SARS-CoV-2

To assess the performance of DRDP for both LFT and NAAT for SARS-CoV-2, paired results after dilution of the same left-over sample in DRDP were compared. Nine Ag tests resulted positive for SARS-CoV-2. All paired PCR tests also tested positive (100% sensitivity).

Out of 27 negative DRDP Ag tests, 25 also tested negative with PCR. Two negative Ag DRDP tests scored weak positive using PCR testing for SARS-CoV-2 with Cq values of 37.47 and 26.59. For the sample with a Cq of 26.59, no officially reported Panbio Ag test result was available. However, because the test cartridge was sent to the lab, a negative result could be read. The diagnostic PCR test for this particular sample was also positive with a Cq value of 23.31. For the second discrepant sample, with a NAAT in DRDP Cq value of 37.47, both diagnostic LFT and NAAT tested negative. However, a Cq value of 37.47 is close to the detection limit of the method.

### 3.4 Influenza A, Influenza B, and RSV nucleic acid test results

Since NAAT was performed using Abbott Alinity m RESP-4-PLEX, also Cq values for Influenza A and B, and RSV were obtained. Unfortunately, no rapid Ag test results were available for any of the aforementioned respiratory viruses. PCR results in IAB (diagnostic PCR test) and DRDP were compared. Five samples tested positive for Influenza A, one sample for Influenza B and none of the samples for RSV. For Influenza A, mean and median Δ Cq values between PCR in IAB and DRDP were 7.12 and 5.25. Δ Cq was 2.28 for Influenza B. A Δ Cq_expected_ of 5.64 for Influenza A (average dilution of 1/50) and 5.87 for Influenza B (average dilution of 1/58) was found taking into account left-over sample volumes for Influenza A or B. Detailed calculations can be found in **Supplementary Data File 1**. As outlined above for SARS-CoV-2, this is in line with the dilution in DRDP that was performed. Overall, the diagnostic results were in agreement with DRDP diluted results for Influenza A and B.

## 4. DISCUSSION

In an ideal scenario for evaluating a novel sample collection buffer, the swab is collected in the buffer itself. However, to minimize patient discomfort, leftover samples from routine rapid antigen testing were used. When comparing the diagnostic LFT results and the LFT results upon dilution in DRDP, we conclude that the use of DRDP universal buffer does not affect rapid antigen test performance since no discrepant results were obtained. Here, the results originated from the same pre-analytical matrix, simplifying data analysis.

In contrast, when comparing the diagnostic NAAT results and NAAT results after dilution in DRDP, a small number of conflicting data points were obtained. Pre-analytically, these results originate from different samplings (as depicted in **Figure 1**) and are therefore independent of each other, complicating interpretation. First, one sample tested strongly positive using the diagnostic NAAT test in IAB (Cq value of 17.69), while the in DRDP diluted counterpart tested negative. Since both the diagnostic LFT for SARS-CoV-2 and the LFT after dilution in DRDP tested negative, a likely explanation can be found in a poor sample quality of the nasopharyngeal swab for diagnostic LFT. Secondly, two samples tested weak positive with NAAT after dilution in DRDP, while the diagnostic NAAT was negative. For one sample with a Cq of 38.26, the diagnostic NAAT tested negative while both diagnostic LFT and DRDP diluted LFT tested positive. This indicates a good agreement between LFT and NAAT after DRDP dilution, while the discrepant result may stem from (1) the pre-analytical phase, or (2) due to the high Cq value (repetition of diagnostic NAAT test might have resulted in a weak positive result). The second sample had a Cq of 37.47 while diagnostic NAAT and LFT tested negative. Repetition of the NAAT in DRDP at day 3 also gave a weak positive result with a Cq of 36.18, confirming the weak positive result. Possibly, while diluting the sample, cross-contamination may have occurred.

Comparing LFT and NAAT results in DRDP, two samples tested negative with LFT, but positive with NAAT. The conflicting result for one sample could be attributed to contamination, as outlined above. For the second sample, an inherent higher sensitivity of NAAT for detection of SARS-CoV-2 likely explains the obtained result.

For some of the discrepant results, mainly when results originate from two different pre-analytical samples, human errors might have played a role. Especially since for molecular testing in DRDP a dilution of the rapid Ag buffer was used, which could explain some discrepancies observed in our dataset. Furthermore, samples were collected in a clinical setting, not in ideal lab circumstances, which resulted in some missing datapoints besides the possibility of human errors.

Since very few data is available for virusPHIX, another universal medium that is commercially available, no comparison was made on this end ^11^.

A limitation of this study is the low number of positive samples included. Additionally, the performance of the buffer on the molecular analyses of other respiratory viruses was only preliminary tested since few positive samples for Influenza A (n = 5) and Influenza B (n = 1) were included. No positive samples for RSV were present in the data set. Unfortunately, no rapid Ag testing was diagnostically performed for Influenza A or B, or RSV at the laboratory of AZ Maria Middelares.

## 5. CONCLUSION

With the data collected in this study, we demonstrated that the DNA/RNA Defend Pro medium is a promising inactivating medium that allows for both reliable Ag testing and PCR analysis for SARS-CoV-2, but also influenza A and influenza B. Additionally, our limited stability results show that RNA is stabilized in DRDP buffer for at least 3 days at room temperature, probably longer. In the future, studies collecting parallel nasal swabs in IAB and DRDP are needed to conclusively demonstrate the usability of DRDP in clinical practice.

## Data Availability

All data produced in the present work are contained in the manuscript

## 6. ACKNOWLEDGEMENTS

The authors wish to acknowledge all laboratory staff of Maria Middelares General Hospital that aided in the reception and processing of the left-over samples. In particular, S.D. would like to specially thank Lies Bouchier, head MLT microbiology, for the practical aid during the study. The ED nurses and doctors are acknowledged for the collection of left-over samples. DNA/RNA Defend Pro medium was provided by InActiv Blue. Figures were created with Biorender.com.

---

**Supplementary Data File 1.**
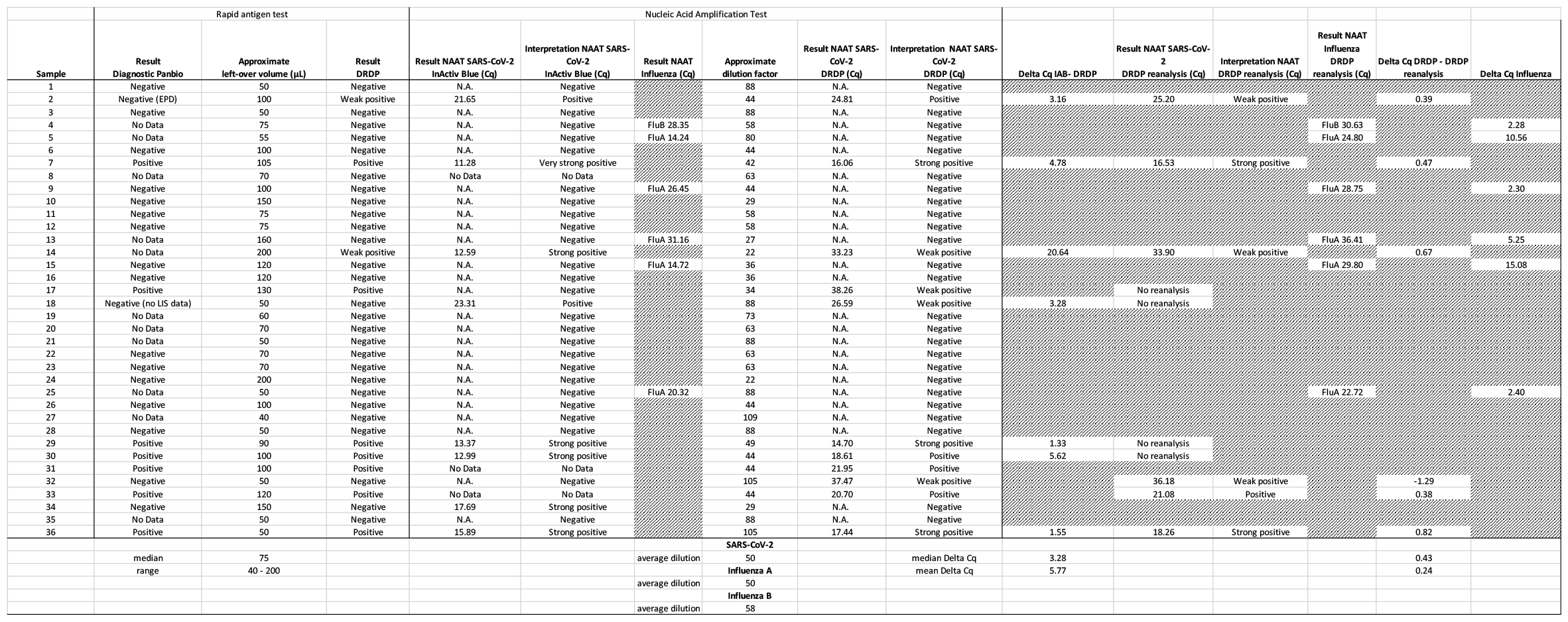

